# Emotional and behavioural phenotypes in young people with neurodevelopmental CNVs

**DOI:** 10.1101/2020.01.28.20019133

**Authors:** Adam C Cunningham, Jeremy Hall, Stewart Einfeld, Michael J Owen, Marianne B M van den Bree

## Abstract

**Background:** A number of disorders caused by copy number variants (CNVs) are associated with a high risk of neurodevelopmental and psychiatric disorders and cognitive impairments (ND-CNVs). Few studies of ND-CNVs have investigated the emotional and behavioural problems that are important outcomes in young people with developmental and intellectual disabilities using appropriate measures.

**Methods:** 322 young people with 13 ND-CNVs across eight loci (mean age:9.79 years, range:6.02-17.91, 66.5% male) took part in the study. Primary carers completed the Developmental Behaviour Checklist (DBC).

**Results:** Sixty-seven percent of individuals with an ND-CNV screened positive for clinically significant difficulties. Young people from families with higher incomes (OR=0.71, CI=0.55 – 0.92, p=.009) were less likely to screen positive. Young people born after prolonged labour (OR=2.87, CI=1.18-8.13, p=.030) were more likely to screen positive. The rate of difficulties differed depending on ND-CNV genotype (Deviance=25.83, p=.011), with the lowest rate in 22q11.2 deletion (46%) and the highest in 1q21.1 deletion (87.5%). Individuals with inherited ND-CNVs had greater difficulties (F=6.54, df=1, p=.012, η_p_^2^=.050), including higher self-absorbed (F=5.01, df=1, p=.027, η_p_^2^=.039) and communication disturbance scores (F=9.13, df=1, p=.003, η_p_^2^=.068). Specific patterns of strengths and weaknesses were found for different ND-CNV genotypes. However, ND-CNV genotype explained no more than 7-16% of variance, depending on subdomain.

**Conclusions:** Behavioural and emotional problems are common in young people with ND-CNVs. The ND-CNV specific patterns we find can provide a basis for more tailored support. More research is needed to better understand the variation in behavioural and emotional problems not accounted for by genotype.

## Introduction

A range of genomic disorders that are caused by sub-microscopic deletions or duplications of various chromosomal regions have been associated with the development of conditions such as ADHD, Autism Spectrum Disorders (ASD), schizophrenia and intellectual disability (ID)(Torres et al., 2015). These changes are referred to as copy number variants (CNVs), as they change the number of copies of genes contained on the affected area of a chromosome. There is strong evidence that CNVs in several chromosomal regions are associated with an increased risk of neurodevelopmental disorder (ND) (referred hereafter as ND-CNVs)(Chawner et al., 2019), however, severity of phenotypic outcomes are variable(Crawford et al., 2018). As technology improves, rates of diagnosis of these conditions are increasing. Therefore, there is a need to improve our understanding of the clinical outcomes associated with these genomic conditions in order to provide optimal counselling and intervention for patients and their families.

Research to date on the mental health profile of individuals with ND-CNVs has highlighted very high rates of neurodevelopmental and psychiatric disorders (Bernier et al., 2016, p. 21; Glassford et al., 2016; Hanson et al., 2014; Schneider et al., 2014; Vermeulen et al., 2017). For many ND-CNVs and psychiatric conditions, such as schizophrenia, the phenotypic presentations are reported to be similar to that seen in non-genotype selective samples (Bassett et al., 2003; Monks et al., 2014). However, for some conditions such as ADHD (Maria Niarchou et al., 2015) or autism spectrum disorder (Angkustsiri et al., 2014), the phenotypic presentation in individuals with ND-CNVs may differ. Furthermore, often the phenotypic presentation is highly complex, with high rates of psychiatric comorbidity (M Niarchou et al., 2014) as well as increased risk of psychiatric disorders in those with motor coordination problems (Cunningham et al., 2018), a history of seizures (Eaton et al., 2019), or sleep disturbances (Moulding et al., 2019). The links between neurocognitive impairments and risk of psychiatric disorder remain to be resolved in individuals with ND-CNVs.

Assessing mental health status in individuals with ID can be difficult. This may particularly be the case for individuals with more severe impairment, where differences in the presentation of psychiatric symptoms have been reported compared to those with mild or moderate ID (Matson & Shoemaker, 2011). Research studies often use standardized assessments of psychiatric disorder that follow the DSM or ICD classification systems. While these assessments allow for common discourse between researchers, as well as comparisons with studies carried out in non-ID populations, they may not adequately assess disorders that present differently or are easily confused with symptoms of ID, such as cognitive slowing (Costello & Bouras, 2006).

ID is common in individuals with ND-CNVs but can range from severe to minimal (Chawner et al., 2019; Hanson et al., 2014; Schneider et al., 2014; Vissers et al., 2016). Despite this, few studies of individuals with ND-CNVs have made use of assessments of psychopathology that are tailored for those with ID (Einfeld et al., 1997; Vermeulen et al., 2017; Wagner et al., 2017). This raises the possibility that previous research might not have assessed certain behaviours that are associated with ID. The Developmental Behaviour Checklist (DBC) (Einfeld & Tonge, 1995) is designed to measure emotional and behavioural problems in young people with ID, and therefore might provide important insights into these behaviours that might have been missed in previous research.

Previous studies of young people with ID have indicated that emotional and behavioural problems can have a significant negative impact on quality of life (Craig et al., 2016), be a common focus of familial stress and be associated with worse mental health in primary carers (Blacher & McIntyre, 2006). Despite their profound impact, there is a lack of research on behavioural problems in young people with ND-CNVs with few studies conducting cross-CNV comparisons (Di Nuovo & Buono, 2011; Vermeulen et al., 2017). Therefore, it is unclear to what extent the wide-ranging phenotypic presentation includes problem domains associated with ID, or if there are differences in the severity of distinct behavioural and emotional problems across individual ND-CNV genotypes. This means that clinicians who see individuals with ND-CNVs often lack information to most effectively counsel these families.

To address the gaps in the literature surrounding behavioural and emotional disturbance profiles in young people with ND-CNVs, we assessed a sample of 322 young people with a range of 13 ND-CNVs across eight loci using the developmental behavioural checklist (DBC) (Einfeld & Tonge, 1995). We investigated the following questions: 1) What proportion of young people with an ND-CNV screen positive for clinically significant behavioural and emotional disturbance as measured using the DBC? 2) What health and pregnancy related variables influence rates of clinically significant difficulties? 3) Are there behavioural and emotional problems that are particularly elevated in young people with ND-CNVs? 4) Are there differences between ND-CNV genotypes in risk for presence of clinically significant difficulties?

## Methods

### Participants

A sample of 322 participants with an ND-CNV (including one of 15q11.2 deletion, 15q13.3 deletion, 15q13.3 duplication, 16p11.2 deletion, 16p11.2 distal deletion, 16p11.2 duplication, 1q21.1 deletion, 1q21 duplication, 22q11.2 deletion, 22q11.2 duplication, 9q34.3 deletion (Kleefstra syndrome), NRXN1 (2p16.3 deletion or TAR (1q21.1) duplication (66.5% male, mean age: 9.79 years, range: 6.02-17.91) who were assessed between September 2011 and November 2018 as part of the ECHO and IMAGINE-ID studies at Cardiff University were included in this study. Data from all instruments used was collected at the same time. Families were recruited through UK Medical Genetics clinics, word of mouth, and charities and support groups for chromosomal disorders including Unique, MaxAppeal! and 22qCrew. Informed and written consent was obtained prior to recruitment from the carers of the children, or the children themselves where appropriate. Recruitment was carried out in agreement with protocols approved by the appropriate university and National Health Service (NHS) ethics and research and development committees. Families were visited at home for phenotyping including cognitive and psychiatric assessment. ND-CNV genotypes were established from medical records as well as in-house genotyping at the Cardiff University MRC Centre for Neuropsychiatric Genetics and Genomics using microarray analysis. Information about the pregnancy, and the children’s medical history, including congenital heart defects, history of epilepsy as well as psychiatric and epilepsy medication use, was collected through primary carer report. The psychiatric and epilepsy medications that were being taken are shown in supplementary table 1.

### The Developmental Behaviour Checklist

Behavioural and emotional problems were assessed using the primary carer version of the Developmental Behaviour Checklist (Einfeld & Tonge, 1995) (DBC). The DBC is designed to asses behavioural and emotional problems in individuals with ID. It has been used in idiopathic ID (Einfeld et al., 2006) as well as in individuals with genetic syndromes associated with ID (Einfeld et al., 1997; Rice et al., 2016; Wagner et al., 2017). The DBC provides a total behaviour problems score (TBPS) as well as scores on five subscales: disruptive/antisocial behaviour (27 items), self-absorbed behaviour (30 items), social relating (10 items), communication disturbance (13 items), and anxiety (9 items). Some items are present in more than one domain. Responses are coded as 0=“not true”, 1=“somewhat true”, 2=“certainly true”. See figure 1 for all items that are included in each of these subscales. Two items, “masturbates or exposes self in public” and “inappropriate sexual behaviour with another” were not included in this study, because the research team deemed these less appropriate to ask the primary carer to complete by questionnaire. Thus, the DBC version we used included total of 94 items. The DBC can also be used to screen for the presence of clinically significant behavioural or emotional difficulties, indicated by a TBPS greater than 45.

**Figure 1.**
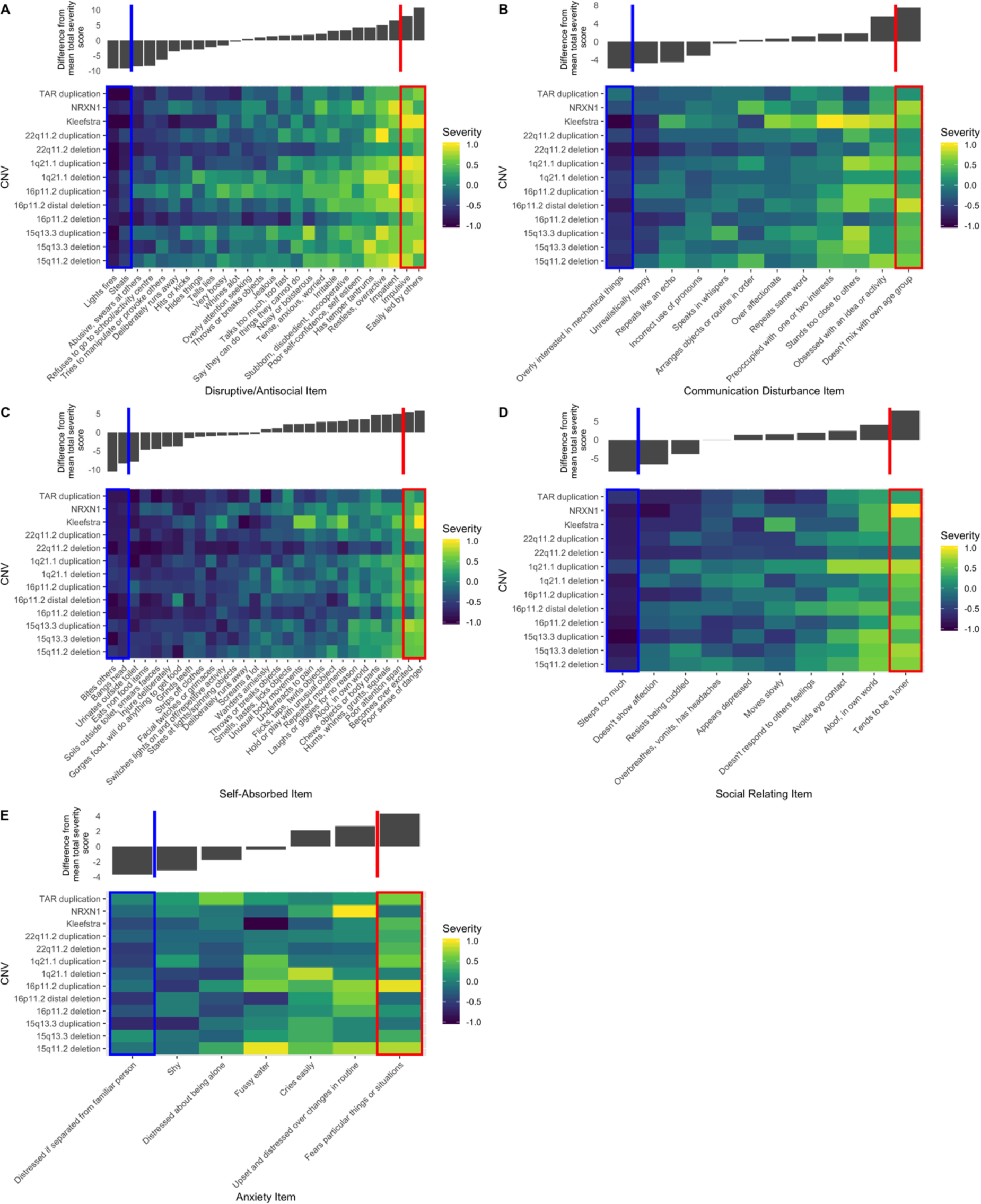
Heat plots of scores for the items constituting the five subscales of the DBC A) disruptive/antisocial, B) communication disturbance C) self-absorbed D) social relating and E) anxiety. Items are ordered on the x-axis corresponding to the total item severity score across the entire ND-CNV sample from lowest on the left to highest on the right. Those items that fall into the highest and lowest 5% of total severity scores are highlighted with red and blue boxes respectively. Bar charts above the heat plots show the deviation from the mean total severity score for each question.

### Statistical Analysis

Statistical analysis was carried out in R version 3.6.1(Development Core Team, 2011). 460/30268 (1.5%) of total DBC responses were missing. These were assumed to be missing at random and imputed using *k-*nearest neighbour imputation with k=5 using the “VIM” package to create a complete dataset (Kowarik & Templ, 2016).

In order to address Aim 1, the percentage of individuals with an ND-CNV displaying clinically significant behavioural difficulties (TBPS >45) was calculated. A logistic regression was used to investigate if age, gender, approximate family income and maternal education level were predictors of the presence of clinically significant behavioural difficulties.

To address aim 2, we used logistic regressions to investigate if health and pregnancy related variables were associated with presence of clinically significant difficulties. We constructed models where presence of clinically significant difficulties was predicted by presence of congenital heart defects; a history of epileptic fits; being part of a multiple birth; whether the mother had fertility treatment during the pregnancy; or a prolonged labour (longer than 36 hours); the baby having spent time in a special care baby unit (SCBU); or in an incubator; or having been born prematurely (before 37 weeks for a single birth, or before 34 weeks for twins). These variables were included in addition to gender, age, maternal education and approximate family income.

In order to address aim 3, we plotted heatmaps of all the items comprising each subdomain to investigate which emotional and behavioural problems impact relatively more or less severely on young people with ND-CNVs (Figure 1 A-E). Each cell represents the deviation from the mean for each question and each ND-CNV genotype. For example, for the disruptive/antisocial subscale (Figure 1A) the average score for each of the 26 items was calculated for each of the 13 ND-CNV genotypes giving a total of 338 values. The mean of these 338 values was calculated, and the deviation from the mean for each ND-CNV and item combination was plotted. Each cell represents the deviation from the overall mean of the 338 values in the plot. To compare between different domains (Figure 1 A-E) these deviations were linearly transformed to fall between 1 and −1 when plotted.

In order to address aim 4, a likelihood ratio test was used to assess if ND-CNV genotype was associated with presence of clinically significant behavioural difficulties after the inclusion of those covariates that were found to be predictors of the presence of clinically significant behavioural difficulties in aim 1 and aim 2 as covariates.

In addition, we used ANCOVAs to test if DBC-TBPS and scores on each of the five subscales differed by ND-CNV genotype after the inclusion of those covariates that were found to be predictors of the presence of clinically significant behavioural difficulties in aim 1 and aim 2. The pattern of severity across the TBPS and subscales was visualised (Fig 2A-F) by plotting the marginal mean TBPS and subscale score for each ND-CNV genotype. These marginal means represent the predicted score after adjusting for those variables that were previously found to be predictors of the presence of clinically significant behavioural difficulties.

**Figure 2.**
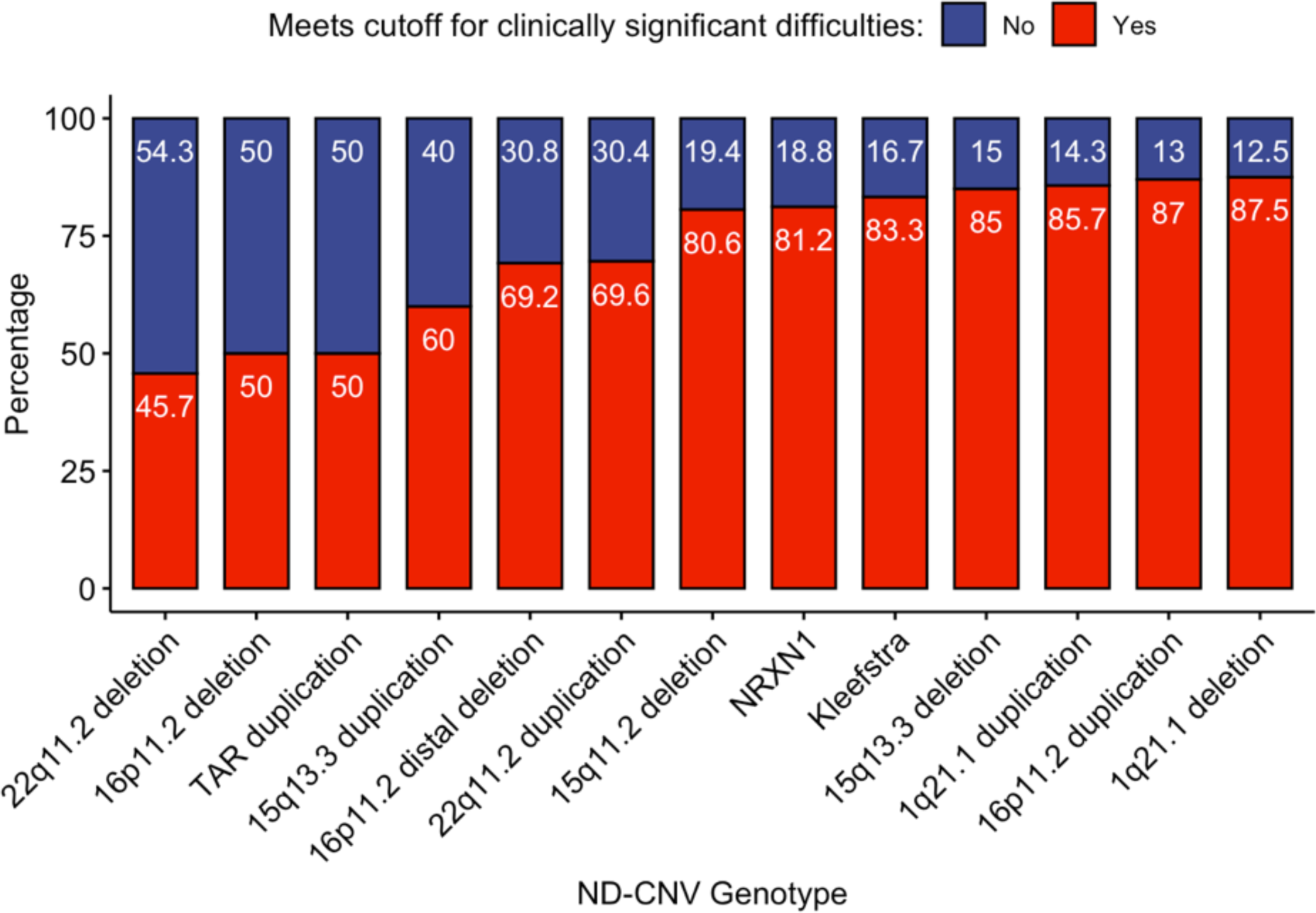
Percentages of individuals screening positive and negative for clinically significant difficulties (DBC total score >45).

We also carried out ANCOVAs to investigate if the type of ND-CNV, (deletion or duplication) or ND-CNV inheritance status (inherited from parent or *de novo*), were associated with TBPS, or scores on each of the five subscales. The association between inheritance status and ND-CNV genotype was tested with a chi-squared test of independence.

As a sensitivity analysis, the procedures outlined for aim 1 and aim 4 were repeated excluding those individuals who were taking psychiatric or epilepsy medication.

In order to test whether there are differences in the severity with which the different ND-CNVs impact on TBPS and the five subscales we used the Kendall’s and Friedman’s chi-square tests, following the same approach as has been previously used to test for quantitative differences in genotype-phenotype associations (Hühn & Piepho, 1994). This allowed us to test the rank concordance (Kendall) and discordance (Friedman) of the ND-CNVs for the different domains.

## Results

Table 1 describes the participants who took part in the study. Participants were aged from 6.012 to 17.91 years (mean age=9.79) and 33.5% were female.

**Table 1.** Descriptive statistics.

|  | Overall (N=322) |
| --- | --- |
| **Maternal education** |  |
| N-Miss | 3 |
| High (University degree and/or other higher postgraduate qualifications) | 86 (27.0%) |
| Middle (A-levels/highers/vocational training) | 127 (39.8%) |
| Low (O-levels/GCSEs) | 79 (24.8%) |
| No School Leaving Exams | 27 (8.5%) |
| **Approximate family income** |  |
| N-Miss | 35 |
| <=£19,999 | 95 (33.1%) |
| £20,000 - £39,999 | 94 (32.8%) |
| £40,000 - £59,999 | 57 (19.9%) |
| £60,000 + | 41 (14.3%) |
| **Child Ethnicity** |  |
| N-Miss | 2 |
| European | 299 (93.4%) |
| Other | 21 (6.6%) |
| **Age of child participants** |  |
| N-Miss | 0 |
| Mean (SD) | 9.794 (3.119) |
| Range | 6.016 - 17.905 |
| **Gender of child participants** |  |
| N-Miss | 0 |
| Female | 108 (33.5%) |
| Male | 214 (66.5%) |
| **Inheritance status** |  |
| N-Miss | 173 (53.7%) |
| De Novo | 79 (24.5%) |
| Inherited | 70 (21.7%) |
| **Full scale IQ (WASI-II)** |  |
| N-Miss | 41 |
| Mean (SD) | 78.811 (14.077) |
| Range | 50.000 - 128.000 |
| **Performance IQ (WASI-II)** |  |
| N-Miss | 36 |
| Mean (SD) | 83.000 (14.695) |
| Range | 53.000 - 132.000 |
| **Verbal IQ (WASI-II)** |  |
| N-Miss | 38 |
| Mean (SD) | 77.986 (14.354) |
| Range | 55.000 - 123.000 |
| **Total behavioural problems score** |  |
| N-Miss | 0 |
| Mean (SD) | 57.186 (27.327) |
| Range | 2.000 - 143.000 |
| **Disruptive/Antisocial subscale** |  |
| N-Miss | 0 |
| Mean (SD) | 19.736 (11.328) |
| Range | 1.000 - 51.000 |
| **Self-Absorbed subscale** |  |
| N-Miss | 0 |
| Mean (SD) | 18.441 (10.742) |
| Range | 0.000 - 52.000 |
| **Communication disturbance subscale** |  |
| N-Miss | 0 |
| Mean (SD) | 8.618 (4.632) |
| Range | 0.000 - 22.000 |
| **Anxiety subscale** |  |
| N-Miss | 0 |
| Mean (SD) | 6.248 (3.567) |
| Range | 0.000 - 14.000 |
| **Social relating subscale** |  |
| N-Miss | 0 |
| Mean (SD) | 4.143 (2.885) |
| Range | 0.000 - 12.000 |
| **Participants taking psychiatric or epilepsy medication** |  |
| N-Miss | 0 |
| No | 286 (88.8%) |
| Yes | 36 (11.2%) |
| **Congenital heart problems** |  |
| N-Miss | 8 |
| No | 247 (78.7%) |
| Yes | 67 (21.3%) |
| **History of epileptic fits** |  |
| N-Miss | 5 |
| No | 260 (82.0%) |
| Yes | 57 (18.0%) |
| **Born prematurely (<37 Weeks)** |  |
| N-Miss | 21 |
| Born prematurely | 149 (49.5%) |
| Normal range | 152 (50.5%) |
| **Child part of multiple birth** |  |
| N-Miss | 14 |
| No | 302 (98.1%) |
| Yes | 6 (1.9%) |
| **Fertility treatment used during pregnancy** |  |
| N-Miss | 19 |
| No | 286 (94.4%) |
| Yes | 17 (5.6%) |
| **Prolonged labour (>36 hours)** |  |
| N-Miss | 23 |
| Normal range | 261 (87.3%) |
| Over 36 hours | 38 (12.7%) |
| **Birth by caesarean section** |  |
| N-Miss | 17 |
| No | 210 (68.9%) |
| Yes | 95 (31.1%) |
| **Child spent time in a special care baby unit** |  |
| N-Miss | 24 |
| No | 236 (79.2%) |
| Yes | 62 (20.8%) |
| **Child spent time in an Incubator** |  |
| N-Miss | 21 |
| No | 240 (79.7%) |
| Yes | 61 (20.3%) |
| WASI-II: Wechsler abbreviated scale of intelligence 2<sup>nd</sup> edition(Wechsler, 1999). |  |

### Aim 1: Proportion of individuals with an ND-CNV screening positive for clinically significant difficulties

215 of 322 (66.8%) individuals with an ND-CNV screened positive for clinically significant difficulties. Logistic regression showed this was predicted by family income (OR=0.71, CI=0.55–0.92, p=.009) with children from families with higher incomes being less likely to experience clinically significant difficulties. Age, gender and maternal education level were not associated with presence of clinically significant difficulties.

### Aim 2: What health and pregnancy related variables influence rates of clinically significant difficulties?

We examined the influence of the following variables on presence of clinically significant difficulties alongside age, gender, maternal education level and approximate family income as covariates: presence of congenital heart defects; a history of epileptic fits; being part of a multiple birth; whether the mother had fertility treatment during the pregnancy; or a prolonged labour (>36 hours); the baby having spent time in a special care baby unit (SCBU); or in an incubator; or having been born prematurely (before 37 weeks for a single birth, or before 34 weeks for twins). Only a prolonged labour was associated with the presence of clinically significant difficulties (OR=2.87, CI=1.18-8.13, p=.030). However, after excluding individuals taking medication, a prolonged labour was no longer associated with clinically significant difficulties.

### Aim 3: Are there behavioural and emotional problems that are particularly elevated in young people with ND-CNVs

In order to investigate what behaviours individuals with ND-CNVs display the most and least difficulties with, we plotted the deviation from the mean for each item and each ND-CNV genotype in a heatmap. We highlighted those items where the sum of severity scores across all ND-CNV groups fell into the highest and lowest 5% of the scores across all questions within a subscale. These results can be found in Figure 1. A-E. The items that have the highest scores are highlighted with red boxes while the lowest scoring items are highlighted with blue boxes.

For the Disruptive/Antisocial subscale, the questions with the highest severity scores were “being easily led by others” and “being impulsive” The lowest severity scores were seen in “lighting fires” and “stealing”.

In the communication disturbance subscale, the items with highest severity were “not mixing well with own age group”, while the lowest severity scores were seen for “being overly interested in mechanical things”.

For the self-absorbed subscale the highest severity scores were seen for “poor sense of danger”, and “becoming over excited”, while the least severe items were “bites others” and “bangs head”.

In the social relating subscale, the most severe item was “tends to be a loner” and the least severe was “sleeps too much”.

Finally, in the anxiety subscale, the most severe item was “fears particular things or situations”, while the least severe was “distressed if separated from a familiar person.”

### Aim 4: Are there differences between ND-CNV genotypes in risk for presence of clinically significant difficulties?

To investigate if the ND-CNV genotypes differed in risk for clinically significant difficulties (based on the total DBC cut-off), we performed a likelihood ratio test comparing models where the presence of difficulties was predicted by family income and prolonged labour with a model where ND-CNV genotype was also included. The addition of ND-CNV genotype improved prediction of the presence of clinically significant difficulties (df=12, Deviance=25.83, p=.011). Fig 2. shows the percentage of individuals displaying clinically significantly difficulties by ND-CNV genotype. Rates of clinically significant difficulties differed across genotypes (χ^2^=41.96, df=12, Fisher’s p<.001). Rates of screening positive ranged from 45.7% (32/70) in 22q11.2 deletion, to 87.5% (14/16) in individuals with 1q21.1 deletion. ND-CNV genotype still improved prediction after excluding individuals taking psychiatric or epilepsy medication (Deviance=22.01, p=.037).

ND-CNV genotype was associated with TBPS and all subscale scores apart from anxiety (Table 2). Table 2 also presents the variance in the TBPS and subdomain scores that is explained by ND-CNV genotype (final column, η_p_^2^). This was found to range from 7.3% for anxiety to 16% of self-absorbed score, along with genotype explaining 15.5% of variance in TBPS. There remained a significant effect of ND-CNV genotype for all scores except anxiety when individuals taking psychiatric or epilepsy medication were excluded (Supplementary table 2).

**Table 2.** Variance explained by ND-CNV genotype on A) DBC total problems score, and B-F) subscale scores, with family income and prolonged labour as covariates.

| **A) DBC total problems** | **F** | **p** | **$\eta_p^2$** |
| --- | --- | --- | --- |
| ND-CNV | 3.83 | 0.000 | 0.155 |
| Family Income | 1.532 | 0.207 | 0.018 |
| Prolonged Labour | 6.265 | 0.013 | 0.024 |
| **B) Disruptive/antisocial** | **F** | **p** | **$\eta_p^2$** |
| ND-CNV | 2.926 | 0.001 | 0.123 |
| Family Income | 1.896 | 0.131 | 0.022 |
| Prolonged Labour | 5.565 | 0.019 | 0.022 |
| **C) Communication disturbance** | **F** | **p** | **$\eta_p^2$** |
| ND-CNV | 2.732 | 0.002 | 0.116 |
| Family Income | 1.182 | 0.317 | 0.014 |
| Prolonged Labour | 2.67 | 0.104 | 0.011 |
| **D) Self-absorbed** | **F** | **p** | **$\eta_p^2$** |
| ND-CNV | 3.974 | 0.000 | 0.160 |
| Family Income | 1.432 | 0.234 | 0.017 |
| Prolonged Labour | 1.832 | 0.177 | 0.007 |
| **E) Social relating** | **F** | **p** | **$\eta_p^2$** |
| ND-CNV | 2.583 | 0.003 | 0.110 |
| Family Income | 1.43 | 0.235 | 0.017 |
| Prolonged Labour | 9.489 | 0.002 | 0.036 |
| **F) Anxiety** | **F** | **p** | **$\eta_p^2$** |
| ND-CNV | 1.644 | 0.080 | 0.073 |
| Family Income | 0.267 | 0.849 | 0.003 |
| Prolonged Labour | 6.769 | 0.010 | 0.026 |

We also used ANCOVAs to investigate if having an inherited (n=79) or *de novo* (n=70) ND-CNV (173 individuals had unknown inheritance) or if the type of genomic change (deletion or duplication) affected TBPS or subscale scores, with family income and prolonged labour as covariates. These tests revealed that having an inherited ND-CNV was associated with greater TBPS (F=6.54, df=1, p=.012, η_p_^2^=.050), self-absorbed score (F=5.01, df=1, p=.027, η_p_^2^=.039), and communication disturbance score (F=9.13, df=1, p=.003, η_p_^2^=.068) but not communication disturbance, social relating, or anxiety score. However, inheritance was also strongly associated with ND-CNV genotype (χ^2^=90.85, df=12, Cramer’s V=0.781, Fisher’s p=<.001), and including both ND-CNV genotype and inheritance status in the model resulted in inheritance no longer being significantly associated with greater problems. Type of genomic change was not associated with TBPS or scores on any of the subscales. Differences in the patterns of severity across DBC subdomains between the ND-CNV genotypes can be seen in Figure 3, which plots marginal mean scores on the TBPS and each of the subscales. These marginal mean scores represent the predicted score after adjusting for family income and prolonged birth status.

**Figure 3.**
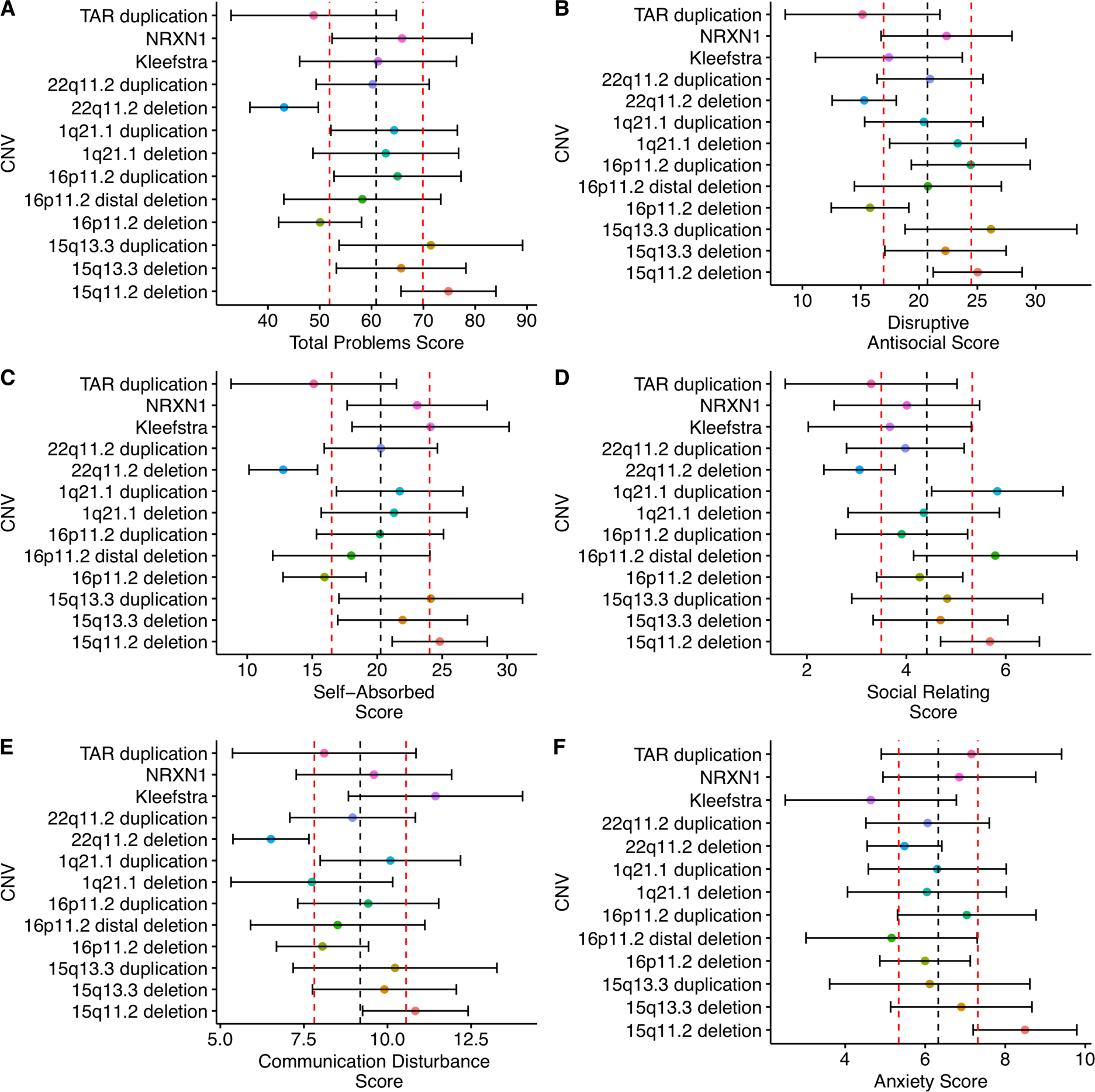
Marginal mean scores for each ND-CNV genotype on A) DBC total problems score and B)–F) the five subscales. The overall group mean for each scale is indicated by the black dashed line, and 1 standard deviation above and below by the dashed red lines. Error bars indicate 95% confidence intervals of the estimated marginal mean. Marginal means are estimated from ANCOVAs including family income and prolonged birth as covariates.

When testing for quantitative differences in these marginal mean scores we found significant concordance in the scores displayed by the young people with ND-CNVs (Kendall χ^2^= 43.23, p<.001). We also observed significant rank discordance in behavioural and emotional problems displayed by the young people with ND-CNVs (Friedman χ^2^=62.89, p<.001). These results suggest that there are both similarities as well as differences in the patterns of behavioural and emotional problems displayed by young people with these ND-CNVs.

## Discussion

This is the first study as far as we are aware that compared phenotypes between a range of different ND-CNV genotypes using a measure of behavioural and emotional problems designed for young people with ID. We demonstrate that behavioural and emotional difficulties are highly prevalent across the ND-CNV genotypes we studied, with 66.8% of young people with ND-CNVs screening positive for clinically significant difficulties. ND-CNV genotype was associated with both screening positive for clinically significant difficulties, as well as TBPS, suggesting that the ND-CNV genotypes display different levels and patterns of difficulties. Furthermore, our rank concordance and discordance analysis indicated that there is evidence of both similarities and dissimilarities in severity, with young people with different ND-CNV genotypes experiencing problems across the subdomains of disruptive/antisocial behaviour, communication disturbances, self-absorbed behaviour, social relating and anxiety. We found that ND-CNV genotype explained approximately 15.5% of variance in TBPS and explained from 7-16% of variance in subscale scores. Type of genomic change (deletion or duplication), did not affect DBC scores. Children with inherited ND-CNVs did display greater overall difficulties, and greater levels of disruptive/antisocial problems, self-absorbed behaviours, and communication disturbance, than children with *de novo* variants. However, inheritance status and ND-CNV genotype were significantly linked, reflecting that rates of inheritance differed between ND-CNV genotypes and we were therefore not able to evaluate the impact of inheritance status independently.

The high prevalence of behavioural and emotional disturbance in this sample of individuals with ND-CNVs agrees with the literature. Previous studies have described the high rates and complex presentations of psychiatric and neurodevelopmental difficulties seen in individuals with ND-CNVs (Chawner et al., 2019; Niarchou et al., 2014; Schneider et al., 2014; Steinman et al., 2016) using standard psychiatric assessments. That we find high rates of psychopathology using a measure that was specifically developed for young people with developmental and intellectual disabilities adds to the robustness of the evidence for high rates of psychopathology in ND-CNVs. We found no effect of gender on rate of clinically significant difficulties which agrees with previous research using the DBC in populations with intellectual disability (Einfeld & Tonge, 1996; Koskentausta & Almqvist, 2004).

We also find that within the ND-CNV sample, there are specific patterns of strengths and weaknesses. Particularly high severity scores were found for “being easily led by others” and “impulsivity”, in the disruptive/antisocial domain, “not mixing well with their own age group”, in the communication disturbance domain, “becoming over excited” and “poor sense of danger” in the self-absorbed domain, “tending to be a loner” in the social relating domain and “fearing particular things or situations” in the anxiety domain. In general, these might suggest that some particular difficulties that individuals with ND-CNVs face are with impulsiveness and social difficulties. These areas may be particularly useful targets for intervention. Difficulties with socialising are likely to contribute to a risk of bullying (Blake et al., 2012) which might compound mental health risk in these individuals (Mayo et al., 2019).

Low severity scores were found for “lighting fires” and “stealing” in the disruptive/antisocial domain, “being overly interested in mechanical things” in the communication disturbance domain, “bites others” and “bangs head” in the self-absorbed domain, “sleeps too much”, in the social relating domain and “distressed if separated from a familiar person” in the anxiety domain, reflecting areas of lower concern in this population.

In the present study, we also find differences in both rates of positive screening for difficulties and on severity scores depending on ND-CNV genotype. This is in addition to the rank discordance that we find in severity of subscales would suggest that there are quantitative differences in the patterns of severity of behavioural and emotional problems across different domains. This means that young people with a specific ND-CNV may be particularly severely affected in one domain such as anxiety, but relatively mildly affected in other domains. However, when taken in combination with the finding of rank concordance in the same data, which suggests that there are similarities between the patterns of severity shown by ND-CNVs across domains, we would conclude that we can find both differences and similarities in behavioural problems across ND-CNV syndromes. This is also highlighted by the low levels of variance explained by ND-CNV genotype in variance in TBPS, suggesting that there are other factors that contribute to psychopathology in these individuals. These could include background genetic risk (potentially measured by a polygenic risk score) (Cleynen et al., 2019) and environmental risk factors, such as bullying or lack of family support. Indeed, we find that approximate family income is predictive of rates of clinically significant difficulties, in agreement with evidence that lower socioeconomic status is associated with greater rates of childhood multimorbidity (Cornish et al., 2013; Johnson et al., 2019). In addition, individuals with inherited ND-CNVs seem to have greater difficulties than individuals with *de novo* variants. It may be that parents who have pathogenic ND-CNVs are less able to cope with their child’s difficulties due to underlying intellectual or mental health symptoms, which increases the child’s risk of developing behavioural problems. This has been shown to occur in depression, with the number of co-occurring problems in mothers with depression predicting psychopathology in their children (Sellers et al., 2013). However, it should be noted that inheritance was strongly associated with ND-CNV genotype and therefore we cannot distinguish between the effect of ND-CNV genotype and inheritance of a variant in the current study.

This is the largest study of its kind to investigate patterns of behavioural and emotional problems across different ND-CNV genotypes. In terms of limitations, there are differences in the size of samples of individuals with different ND-CNVs. Ascertainment bias may affect our results, as developmental delay is a major reason for referral for genetic testing in the UK. This might be highlighted by our findings of individuals with 15q11.2 deletion having some of the most severe scores on the DBC, despite continued debate over the clinical significance of this ND-CNV genotype (Butler, 2017), along with the relatively lower problems displayed by individuals with 22q11.2 deletion are more likely to be referred to genetics clinics for medical issues. It may be that the full range of outcomes (including no or mild phenotypic effects) are not captured in this study. However, even if we are predominantly including those who are also more likely to have a higher load of genetic background and environmental risk factors, it is still important to better understand the difficulties faced by this group of patients, as they make up a significant proportion of those presenting for clinical genetic testing and care following a CNV diagnosis. Clinicians such as genetic counsellors or paediatricians often lack complete information on the likely outcomes of patients with an ND-CNV in their care and are therefore unable to provide accurate counselling or advice to patients or their families. Improving our knowledge and understanding of behavioural and emotional risk is therefore key to improving counselling provision and access to appropriate services. This is particularly relevant as rates of patients presenting for genetic screening will increase as technology to detect chromosomal aberrations continues to improve.

### Clinical implications

Immediate clinical implications of these findings should be increased vigilance for behavioural and emotional problems in individuals with ND-CNVs. If ID is suspected, then psychiatric assessments that are designed to be used in individuals with ID should be applied. Particular difficulties in individuals with ND-CNVs may be impulsiveness and poor social skills. However, it would be best for every child to complete a DBC so that interventions can be targeted at an individual level. In addition, patients who display both ID and behavioural or emotional problems in the absence of any physical injury that could cause of ID, are potential candidates for genetic screening which may reveal contributory genetic variants. Individuals with inherited variants, and those from lower income backgrounds may be at greater risk for clinically significant difficulties.

## Conclusions

Behavioural and emotional difficulties are common in individuals with ND-CNVs. These problems may show specific patterns depending on ND-CNV genotype, but individuals with ND-CNVs show particular problems with social relating, attention, impulsivity and fearing particular things or situations. Inclusion of measures like the DBC which are adapted for ID in research involving young people with ND-CNVs is recommended in future research.

## Data Availability

Data Available on request

## Notes

**Conflicts of Interest:** None

### Competing Interest Statement

The authors have declared no competing interest.

### Funding Statement

Funding: This research was funded by MRC grant Intellectual Disability and Mental Health: Assessing Genomic Impact on Neurodevelopment (IMAGINE) (MR/N022572/1 and MR/L011166/1; JH, MvdB and MO), a Wellcome Trust Strategic Award ‘Defining Endophenotypes From Integrated Neurosciences (DEFINE) (503147 MO and JH), Medical Research Council Programme Grant (G0800509; MO), the National Institute of Mental Health (NIMH 5UO1MH101724; MvdB and MO), a Wellcome Trust Institutional Strategic Support Fund (ISSF) award (MvdB), the Waterloo Foundation (918-1234; MvdB), and an early career fellowship from the Waterloo Foundation Changing Minds programme awarded to AC, the Baily Thomas Charitable Fund (2315/1; MvdB and TRUST/VC/AC/SG/5196-8188; MvdB) and Health & Care Research Wales (Welsh Government, 507556).

